# Association between zidovudine and adverse pregnancy outcomes/congenital malformations: A pharmacovigilance study using FAERS data

**DOI:** 10.1101/2025.08.19.25334037

**Authors:** Zhongxiang Zhang, Xinyun Du, Liuyi Ren, Long He, Xuping Yang, Qiaoying Li, Kun Tu, Shurong Wang, Jie Zhou, Yilan Huang

## Abstract

**Background:** Zidovudine (AZT), a key antiretroviral drug used for HIV treatment and preventing mother-to-child transmission, has insufficient post-marketing pharmacovigilance regarding pregnancy outcomes and congenital disorders.

**Methods:** This pharmacovigilance study analyzed adverse event (AE) reports associated with zidovudine from the U.S. FDA Adverse Event Reporting System (FAERS) database (Q1 2004–Q4 2024) to assess its safety in pregnancy. Four statistical methods were used: Reporting Odds Ratio (ROR), Proportional Reporting Ratio (PRR), Bayesian Confidence Propagation Neural Network (BCPNN), and Multi-item Gamma Poisson Shrinker (MGPS).

**Results:** A total of 2,931 case reports (12,586 adverse events) involving zidovudine as the primary suspect drug were analyzed, with 802 cases reported from pregnant individuals. Disproportionality analysis revealed significant associations with blood and lymphatic disorders, pregnancy and perinatal complications, congenital and genetic disorders, as well as hepatobiliary conditions. Notable signals included preterm birth [ROR(95% CI) = 32.61 (28.36, 37.49)], low birth weight [ROR(95% CI) = 20.44 (14.51, 28.78)], and congenital anomalies [ROR(95% CI) = 26.54 (19.58, 35.96)]. Additionally, new unlabeled signals, such as acquired lipodystrophy and atrial septal defects, were identified.

**Conclusion:** The findings from this pharmacovigilance study enhance the post-marketing safety monitoring of zidovudine, inform clinical decisions in pregnant populations, and highlight the need for targeted surveillance and risk-benefit assessment. Future research should validate novel signals and compare different antiretroviral regimens in pregnancy.

## 1. Introduction

The human immunodeficiency virus (HIV) continues to pose a significant burden on global public health, affecting many countries. As of 2024, 40.8 million people worldwide are living with HIV, 1.3 million people were newly infected with HIV, and 630,000 people died from AIDS-related illnesses(1). Annually, approximately 1.3 million HIV-positive individuals become pregnant, with around 1.1 million (85%) receiving ART during their pregnancies(2). Access to antiretroviral therapy (ART) for HIV-positive pregnant and lactating women remains a cornerstone in the global strategy to reduce vertical transmission of HIV(3).

Zidovudine (AZT), the first antiretroviral drug approved for treating HIV/AIDS, falls under the nucleoside reverse transcriptase inhibitor (NRTI) class(4). It functions by being phosphorylated to zidovudine triphosphate by cytosolic thymidine kinase within virus-positive cells. This metabolite selectively inhibits the HIV reverse transcriptase enzyme, halting HIV strand synthesis and thus preventing viral replication(5). Historically, zidovudine was a first-line agent for preventing mother - to - child transmission (MTCT). However, in most regions, it has been supplanted by tenofovir - based regimens. Despite this, zidovudine still sees use in resource - limited settings or for short - term perinatal prophylaxis(6, 7). Given its continued application in specific scenarios, a comprehensive understanding of its safety profile, especially regarding potential adverse pregnancy outcomes, is crucial for healthcare providers.

Previous research has explored the link between zidovudine and pregnancy - related disorders. A systematic review and meta - analysis involving 124,478 pregnant women living with HIV (WLHIV) demonstrated that ART containing zidovudine was associated with an elevated risk of adverse perinatal outcomes, such as very preterm labor, compared to zidovudine - free ART(8). Data from the Antiretroviral Pregnancy Registry (APR), which included 12,780 singleton pregnancies with intrauterine zidovudine exposure from January 1, 1989, to July 31, 2013, revealed that zidovudine based regimens were significantly associated with a higher risk of low birth weight (LBW) compared to zidovudine free antiretroviral (ARV) regimens(9). Additionally, other studies reported a significant correlation between zidovudine use during pregnancy and hypertrophied fetal hearts, along with indications of increased mitochondrial numbers in HIV - exposed uninfected (HEU) fetuses(10, 11).

Collectively, existing evidence strongly suggests a correlation between zidovudine and the risk of pregnancy-related disorders. However, the current data do not definitively establish this relationship. To further validate previous findings and comprehensively assess zidovudine’s safety, this study leverages the Food and Drug Administration Adverse Event Reporting System (FAERS) database. By applying a range of statistical methods, including Reporting Odds Ratio (ROR), Proportional Reporting Ratio (PRR), Bayesian Confidence Propagation Neural Network (BCPNN), and the Multi-item Gamma Poisson Shrinker (MGPS) method (with Empirical Bayesian Geometric Mean [EBGM] as its core metric), we conduct a detailed analysis of individual drug reports in the FAERS database(12–15).

## 2. Materials and methods

### 2.1 Data source

Data were obtained from the publicly available FAERS database, covering the period from Q1 2004 to Q4 2024. The FAERS is a comprehensive database that includes reports of adverse events (AEs), medication errors, and product quality complaints leading to AEs, with data contributed by healthcare professionals (e.g., physicians, pharmacists), manufacturers, and other individuals(16, 17). For this study, we focused on reports involving pregnant individuals, identified through MedDRA terms related to pregnancy (e.g., "Pregnancy exposure," "Fetal exposure during pregnancy") to ensure accuracy in defining the target population.

Adverse events and medication errors in the FAERS database are coded using terminology from the Medical Dictionary for Regulatory Activities (MedDRA). In the present study, we employed the latest version of the MedDRA lexicon (MedDRA 27.1) for coding adverse event names, with a specific focus on System Organ Classes (SOCs) and Standardized MedDRA Queries (SMQs) pertinent to pregnancy (e.g., "Pregnancy, puerperium and perinatal conditions") and congenital disorders (e.g., "Congenital, familial and genetic disorders").

The date for obtaining data for research purposes is January 31, 2025. During or after the data collection period, the author is unable to obtain any information that can identify individual participants because all reports in the FAERS database have undergone strict anonymization, removing any information that can identify the identity of an individual, including names, ID numbers, contact information, and other private data. This means that throughout the entire research process, the author cannot trace back to any individual participant using the obtained data, effectively safeguarding the privacy and personal information security of the participants, and also complying with the ethical norms and privacy protection requirements for data use in retrospective studies.

Research using data obtained from the FAERS database does not require ethical review. This is because the data has been rigorously anonymized, does not contain any personally identifiable information, and is a publicly available resource for scientific research. This study is a retrospective observational analysis that does not involve any individual intervention or harm and complies with the exemption clause for secondary use of public databases in international ethical guidelines.

### 2.2 Data cleansing

Due to the spontaneous submission of data to the database, there are instances of duplicate reports, withdrawn reports, and deleted reports. Consequently, the FDA’s official guidance document outlines specific rules for data de-duplication and provides a list of reports that should be deleted. For this study, we strictly followed the FDA’s guidance document, which is available on their official website for data cleaning procedures (Fig 1).

**Fig 1.**
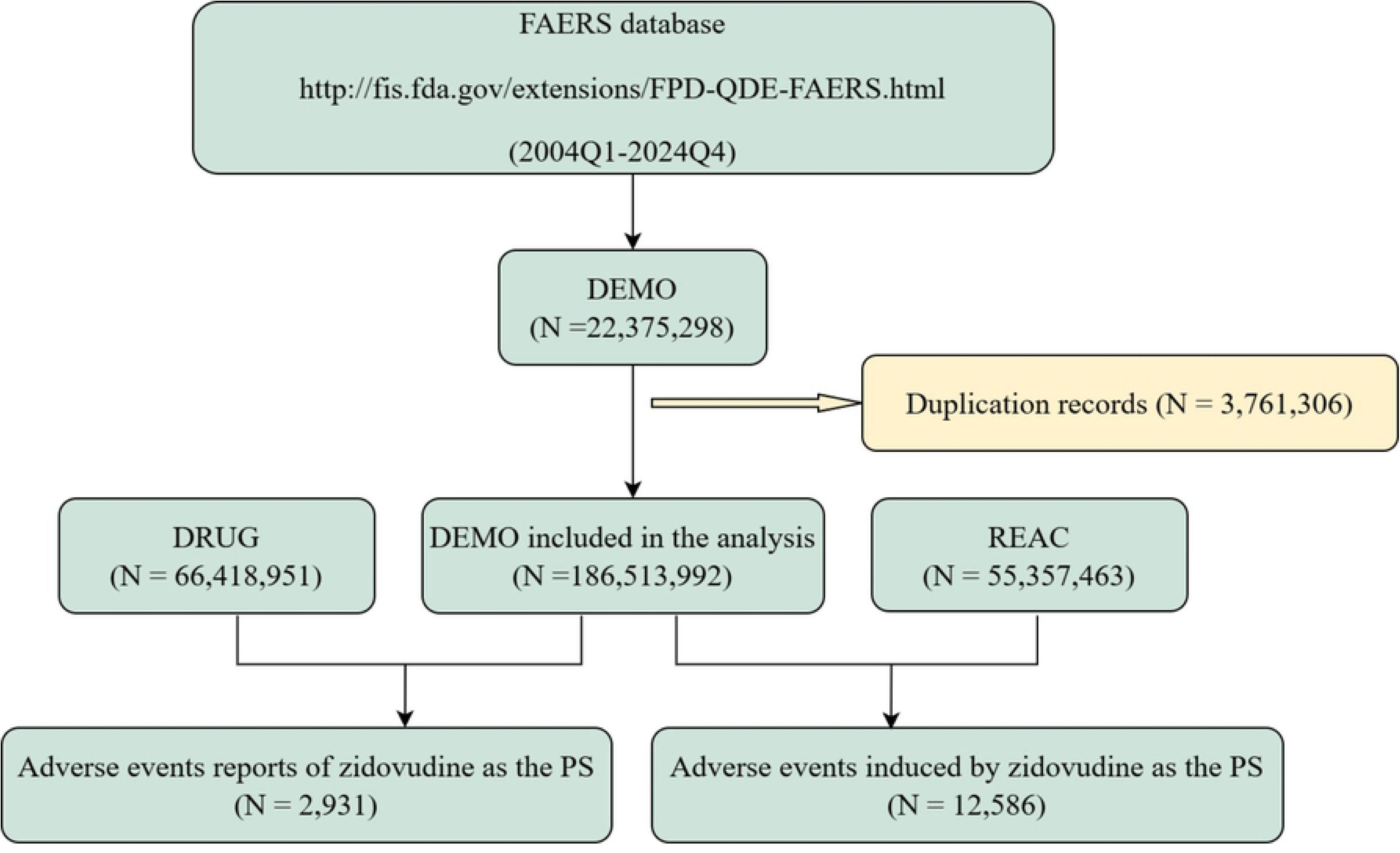
The flow diagram of selecting tagset Drug AEs in the overa.

The data cleaning process involved several key steps. First, in accordance with the FDA’s recommended method for removing duplicate reports, the PRIMARYID, CASEID, and FDA_DT fields from the DEMO table were utilized. Reports with the same CASEID were organized by CASEID, FDA_DT, and PRIMARYID, retaining those with the most recent FDA_DT values. In cases where reports had identical CASEID and FDA_DT values, the report with the highest PRIMARYID value was preserved. Second, a list of deleted reports has been provided in each quarterly packet since Q1 2019. Following data de-weighting, reports were eliminated based on their CASEID as listed in the deletion records.

### 2.3 Screening of the target drug population

Each patient report will feature a unique Primary Suspect Drug (PS) in the database, with only the patient’s PS being considered when determining the target drug use population. If a patient’s Primary Suspect Drug corresponds to the target drug under investigation in the background database, that patient will be included in the target drug population; otherwise, they will be categorized into the other drug population.

The WHO DRUG Dictionary (September 2024 edition) was utilized to standardize all drug names within the database, and the standardized generic names were employed for screening purposes.

### 2.4 Statistical analysis

Based on MedDRA terminology, disproportionality analyses of adverse events from the FAERS were conducted at the System Organ Class, Preferred Term, and Standardised MedDRA Query levels, with a specific focus on SMQs relevant to pregnancy outcomes (e.g., "Pregnancy, labour and delivery complications and risk factors") and congenital disorders (e.g., "Congenital, familial and genetic disorders"). Disproportionality analysis, a well-established approach for detecting safety signals from individual case safety reports, was central to our methodology(18). The analytical methods employed included ROR, PRR, BCPNN, and MGPS, with detailed formulas provided in S1 table.

Positive AE signals were defined by meeting the thresholds of all four methods: ROR (number of reports ≥ 3, lower 95% confidence interval [CI] > 1); PRR (χ ² ≥ 4, lower 95% CI > 1); BCPNN (information component lower 95% CI [IC025] > 0); and MGPS (empirical Bayesian geometric mean lower 95% CI [EBGM05] > 2).

### 2.5 Statistical analysis software

This study employed SAS 9.4 for statistical analysis, a software recommended by the FDA for mining the FAERS database. Data analysis was further supported by Navicate (version 16), Microsoft Excel (version 2021), and SPSS (version 27.0.1). Visualization of the results was conducted using GraphPad (version 9.5).

## 3. Results

### 3.1 General characteristics

The analysis encompassed a total of 18,613,992 background patients, with 55,357,463 AEs reported over 84 quarters spanning from Q1 2004 to Q4 2024. Within this dataset, 2,931 patients were identified as part of the target drug population (zidovudine as the primary suspect drug), corresponding to 12,586 AEs. All statistics were presented on a patient-level basis, meaning each patient was counted only once regardless of the number of concurrent AEs they experienced. The analysis included a comprehensive assessment of various indicators, such as sex, age, reporter occupation, country of reporting, and year of reporting, with detailed breakdowns provided in Table 1.

**Table 1.** Summary of demographic information on the population of the drug zidovudine.

|  | Characteristics | CaseNumber,n | Proportions (%) |
| --- | --- | --- | --- |
| Gender | Female | 857 | 29.24 |
|  | Male | 958 | 32.69 |
|  | Not Specified | 1116 | 38.08 |
| Age (years) | <18 | 502 | 17.13 |
|  | 18~44 | 508 | 17.33 |
|  | 45~65 | 283 | 9.66 |
|  | >65 | 16 | 0.55 |
|  | Not Specified | 1622 | 55.34 |
| Indications (top five) | HIV infection | 1409 | 48.07 |
|  | Antiviral prophylaxis | 296 | 10.1 |
|  | Prophylaxis against HIV infection | 194 | 6.62 |
|  | Antiretroviral therapy | 167 | 5.7 |
|  | Product used for unknown indication | 135 | 4.61 |
| Occupation of reporters | Consumer | 587 | 20.03 |
|  | Lawyer | 2 | 0.07 |
|  | Other health-professional | 1093 | 37.29 |
|  | Pharmacist | 331 | 11.29 |
|  | Physician | 593 | 20.23 |
|  | Not Specified | 325 | 11.09 |
| Reported countries (top five) | United States of America | 796 | 27.16 |
|  | France | 404 | 13.78 |
|  | United Kiongdom | 180 | 6.14 |
|  | India | 131 | 4.47 |
|  | Japan | 101 | 3.45 |
| Outcomes |  |  |  |
|  | Life-Threatening | 116 | 3.96 |
|  | Hospitalization - Initial or Prolonged | 708 | 24.16 |
|  | Disability | 52 | 1.77 |
|  | Death | 362 | 12.35 |
|  | Congenital Anomaly | 360 | 12.28 |
|  | Required Intervention to Prevent Permanent Impairment/Damage | 37 | 1.26 |
|  | Other | 1895 | 64.65 |
| Reported years |  |  |  |
|  | 2004 | 148 | 5.05 |
|  | 2005 | 149 | 5.08 |
|  | 2006 | 257 | 8.77 |
|  | 2007 | 106 | 3.62 |
|  | 2008 | 143 | 4.88 |
|  | 2009 | 205 | 6.99 |
|  | 2010 | 139 | 4.74 |
|  | 2011 | 140 | 4.78 |
|  | 2012 | 71 | 2.42 |
|  | 2013 | 99 | 3.38 |
|  | 2014 | 119 | 4.06 |
|  | 2015 | 175 | 5.97 |
|  | 2016 | 361 | 12.32 |
|  | 2017 | 188 | 6.41 |
|  | 2018 | 101 | 3.45 |
|  | 2019 | 74 | 2.52 |
|  | 2020 | 110 | 3.75 |
|  | 2021 | 97 | 3.31 |
|  | 2022 | 113 | 3.86 |
|  | 2023 | 90 | 3.07 |
|  | 2024 | 46 | 1.57 |

A key focus of this study was the subset of patients with confirmed pregnancy status, identified through MedDRA terms (e.g., "Pregnancy exposure," "Fetal exposure during pregnancy") and supplementary clinical records (e.g., gestational age documentation, delivery notes), totaling 802 patients. This pregnancy-specific subset was central to evaluating zidovudine’s safety profile in the context of pregnancy outcomes and congenital disorders.

Notably, in the overall target population, a higher proportion of AEs was reported among patients younger than 18 years (17.13%) and those aged 18 to 44 years (17.33%). It should be clarified that the under-18 group is not part of the "pregnant population" in this study, as the latter was strictly defined via MedDRA terms (e.g., "Pregnancy exposure") and supplementary clinical records (802 cases total). The under-18 group was analyzed separately to reflect zidovudine’s safety profile in adolescents and children. This distribution suggests that zidovudine-associated AEs may be more prevalent in adolescent and young adult populations, requiring increased vigilance. Particularly, the 18 to 44 years range includes the primary reproductive cohort. Meanwhile, indications show HIV infection (48.07%) as the top occurrence, followed by antiviral prophylaxis (10.1%), aligning with zidovudine’s established role in HIV prevention and treatment and reinforcing the dataset’s clinical relevance.

In terms of geographic distribution, the United States contributed the largest share of AE reports (27.16%), which may reflect higher reporting rates or greater clinical usage in this region. Among the reported outcomes, hospitalization (initial or prolonged) was the most common (708 cases, 24.16%), underscoring the potential severity of some zidovudine-associated AEs. Other notable severe outcomes included death (362 cases, 12.35%) and congenital anomalies (360 cases, 12.28%), with the latter being of particular interest for the pregnancy-specific analyses. The number of reports varied across years, with 2016 recording the highest count (361 cases, 12.32%), a trend that may correlate with increased clinical attention to zidovudine’s safety profile during that period.

### 3.2 Disproportionality analysis

The signal intensity of zidovudine at the SOC level is illustrated in Fig 2. To identify statistically significant SOC signals, we applied four algorithms for disproportionality analysis, with the results presented in Table 2. Our analysis confirmed that the eligible SOCs with significant signals included Blood and lymphatic system disorders, Pregnancy, puerperium and perinatal conditions, Congenital, familial and genetic disorders, and Hepatobiliary disorders—all of which met the threshold criteria for all four methods (ROR, PRR, BCPNN, and MGPS).

**Fig 2.**
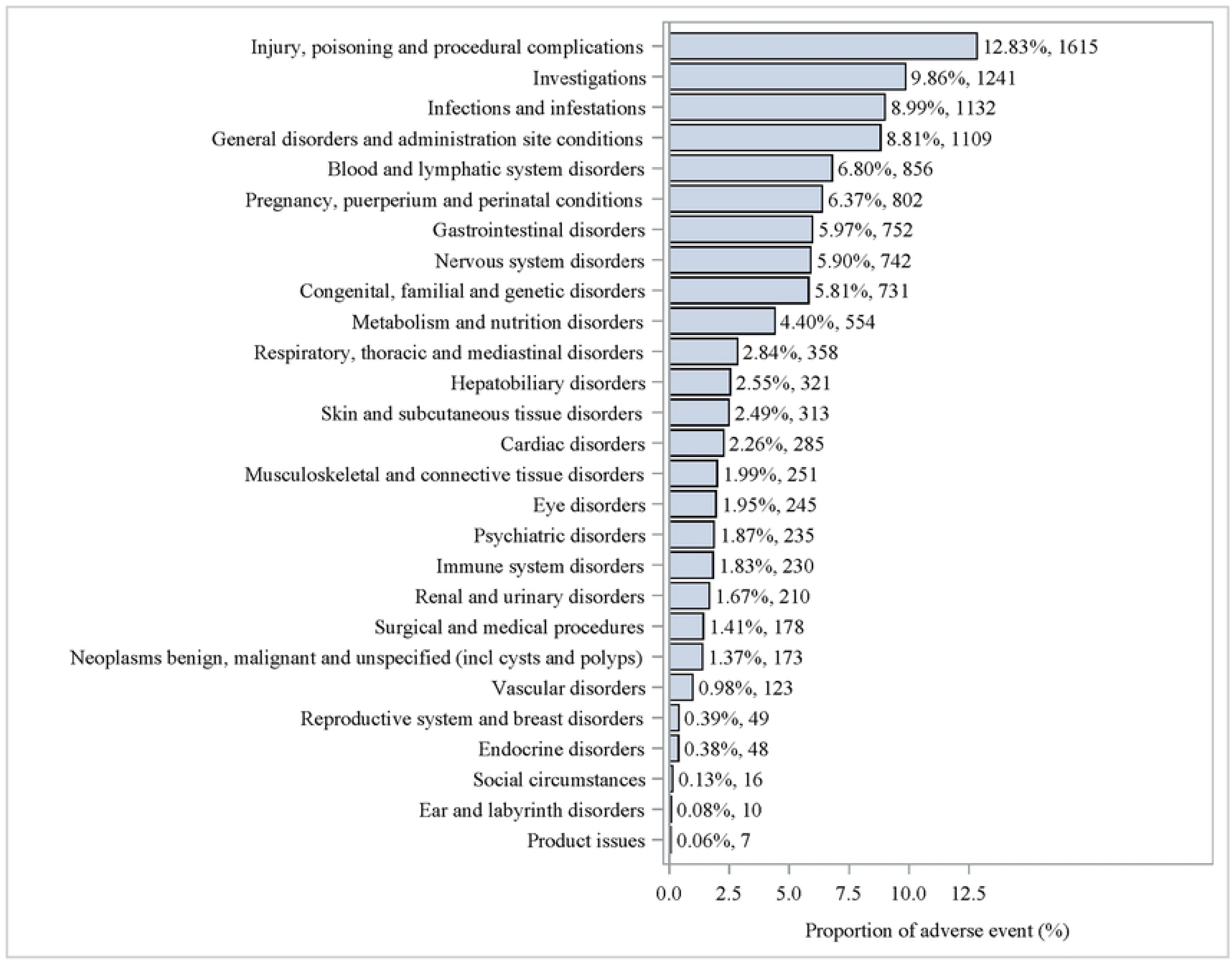
Proportion of adverse events by socs.

**Table 2.** SOC Hierarchy Signal Calculation Results.

| SOC | The report number | ROR (95% CI) | PRR (Chi_Square) | IC (IC025) | EBGM (EBGM05) |
| --- | --- | --- | --- | --- | --- |
| Blood and lymphatic system disorders | 856 | 4.25 (3.97,4.56) | 4.03 (1982.55) | 2.01 (1.90) | 4.03 (3.76) |
| Pregnancy, puerperium and perinatal conditions | 802 | 15.88 (14.78,17.06) | 14.93 (10432.3) | 3.90 (3.77) | 14.88 (13.85) |
| Congenital, familial and genetic disorders | 731 | 20.54 (19.06,22.13) | 19.40 (12741.3) | 4.27 (4.13) | 19.32 (17.93) |
| Hepatobiliary disorders | 321 | 2.83 (2.53,3.16) | 2.78 (369.08) | 1.47 (1.30) | 2.78 (2.49) |

Among these, Blood and lymphatic system disorders, involving 856 cases from the overall target population, showed an overall signal intensity characterized by [ROR (95% CI) = 4.25 (3.97, 4.56); PRR (95% CI) = 4.03 (3.78, 4.30); PRR (Chi_Square) = 4.03 (1982.55); IC (IC025) = 2.01 (1.90); EBGM (EBGM05) = 4.03 (3.76)]. In contrast, the Pregnancy, puerperium and perinatal conditions SOC, which focused exclusively on the 802 confirmed pregnant individuals, included 802 cases with a notably high signal intensity:[ROR (95% CI) = 15.88 (14.78, 17.06); PRR (95% CI) = 14.93 (13.96, 15.97); PRR (Chi_Square) = 14.93 (10432.3); IC (IC025) = 3.90 (3.77); EBGM (EBGM05) = 14.88 (13.85)]. Congenital, familial and genetic disorders, a key focus for pregnancy-related safety, included 731 cases with a signal significance of [ROR (95% CI) = 20.54 (19.06, 22.13); PRR (95% CI) = 19.40 (18.08, 20.82); PRR (Chi_Square) = 19.40 (12741.3); IC (IC025) = 4.27 (4.13); EBGM (EBGM05) = 19.32 (17.93)]. Hepatobiliary disorders, with the lowest number of cases at 321, had a signal intensity of [ROR (95% CI) = 2.83 (2.53, 3.16); PRR (95% CI) = 2.78 (2.50, 3.10); PRR (Chi_Square) = 2.78 (369.08); IC (IC025) = 1.47 (1.30); EBGM (EBGM05) = 2.78 (2.49)]. These findings highlight specific areas where zidovudine-related adverse events are most frequently reported, with the pregnancy-specific SOCs showing particular relevance to the pregnant population.

We further conducted a disproportionality analysis at the Standardized MedDRA Query (SMQ) level, with results presented in Table 3. Among the analyzed outcomes, those specifically relevant to pregnancy and congenital disorders included: "Complications and Risk Factors of Pregnancy, Labor, and Delivery (other than Miscarriage and Stillbirth)," "Congenital, Familial, and Hereditary Diseases," "Fetal disorders," "Termination of pregnancy and risk of abortion," "Congenital, familial, neonatal, and genetic disorders of the liver," and "Congenital and neonatal arrhythmias."

**Table 3.**
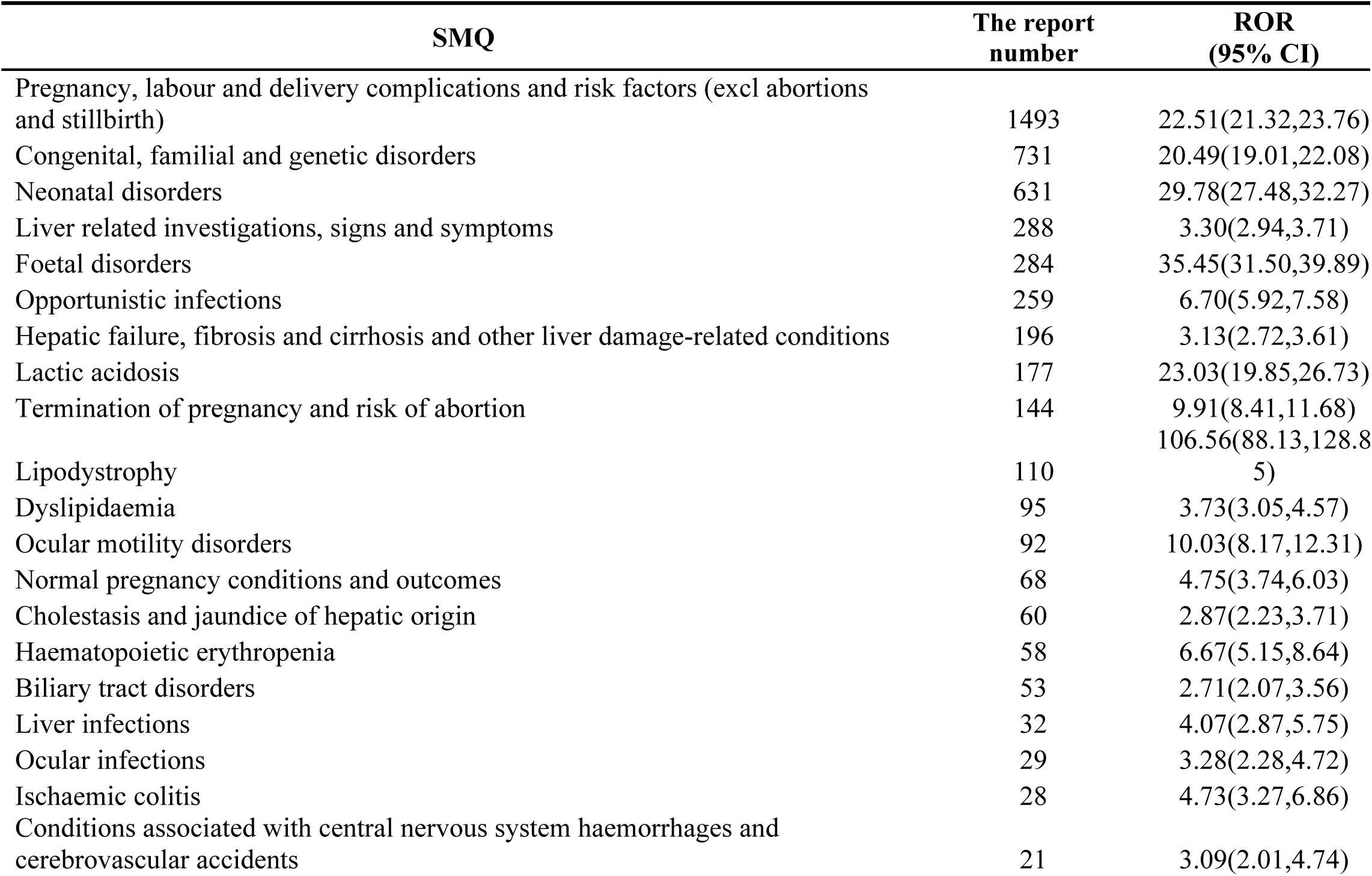
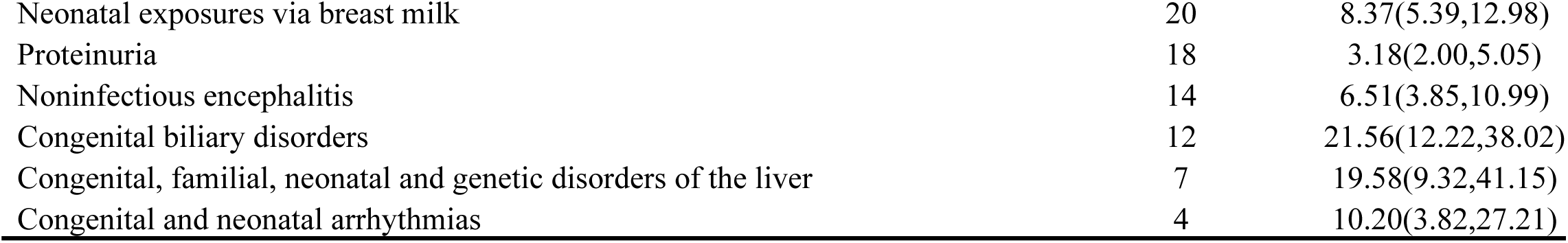
SMQ Hierarchy Signal Calculation Results.

| SMQ | The report number | ROR (95% CI) |
| --- | --- | --- |
| Pregnancy, labour and delivery complications and risk factors (excl abortions and stillbirth) | 1493 | 22.51(21.32,23.76) |
| Congenital, familial and genetic disorders | 731 | 20.49(19.01,22.08) |
| Neonatal disorders | 631 | 29.78(27.48,32.27) |
| Liver related investigations, signs and symptoms | 288 | 3.30(2.94,3.71) |
| Foetal disorders | 284 | 35.45(31.50,39.89) |
| Opportunistic infections | 259 | 6.70(5.92,7.58) |
| Hepatic failure, fibrosis and cirrhosis and other liver damage-related conditions | 196 | 3.13(2.72,3.61) |
| Lactic acidosis | 177 | 23.03(19.85,26.73) |
| Termination of pregnancy and risk of abortion | 144 | 9.91(8.41,11.68) |
| Lipodystrophy | 110 | 106.56(88.13,128.85) |
| Dyslipidaemia | 95 | 3.73(3.05,4.57) |
| Ocular motility disorders | 92 | 10.03(8.17,12.31) |
| Normal pregnancy conditions and outcomes | 68 | 4.75(3.74,6.03) |
| Cholestasis and jaundice of hepatic origin | 60 | 2.87(2.23,3.71) |
| Haematopoietic erythropenia | 58 | 6.67(5.15,8.64) |
| Biliary tract disorders | 53 | 2.71(2.07,3.56) |
| Liver infections | 32 | 4.07(2.87,5.75) |
| Ocular infections | 29 | 3.28(2.28,4.72) |
| Ischaemic colitis | 28 | 4.73(3.27,6.86) |
| Conditions associated with central nervous system haemorrhages and cerebrovascular accidents | 21 | 3.09(2.01,4.74) |
| Neonatal exposures via breast milk | 20 | 8.37(5.39,12.98) |
| Proteinuria | 18 | 3.18(2.00,5.05) |
| Noninfectious encephalitis | 14 | 6.51(3.85,10.99) |
| Congenital biliary disorders | 12 | 21.56(12.22,38.02) |
| Congenital, familial, neonatal and genetic disorders of the liver | 7 | 19.58(9.32,41.15) |
| Congenital and neonatal arrhythmias | 4 | 10.20(3.82,27.21) |

The category "Complications and Risk Factors of Pregnancy, Labor, and Delivery (other than Miscarriage and Stillbirth)" had the highest number of reported AEs, with 1,493 cases, and a disproportionality result of [ROR (95% CI) = 22.51 (21.32, 23.76)]. The second most frequent category was "Congenital, Familial, and Hereditary Diseases," with 731 cases and a signal strength of [ROR (95% CI) = 20.49 (19.01, 22.08)]. "Fetal disorders," which directly relates to in utero exposure outcomes, included 284 cases with a notably high ROR: [ROR (95% CI) = 35.45 (31.50, 39.89)]. "Termination of pregnancy and risk of abortion" recorded 144 cases, with results of [ROR (95% CI) = 9.91 (8.41, 11.68)]. The categories with the lowest number of AE reports were "Congenital, familial, neonatal, and genetic disorders of the liver" (7 cases) and "Congenital and neonatal arrhythmias" (4 cases), yielding ROR values of [19.58 (9.32, 41.15)] and [10.20 (3.82, 27.21)], respectively—though these should be interpreted cautiously due to the small sample size.

At the Preferred Term (PT) level, a total of 382 AE signals were identified, with the largest number of signals associated with Congenital, familial and genetic disorders (n = 65), followed by Pregnancy, puerperium and perinatal conditions (n = 33), which can be seen in S2 table. The top 50 AEs, categorized by their frequency following zidovudine administration, are illustrated in Fig 3.

**Fig 3.**
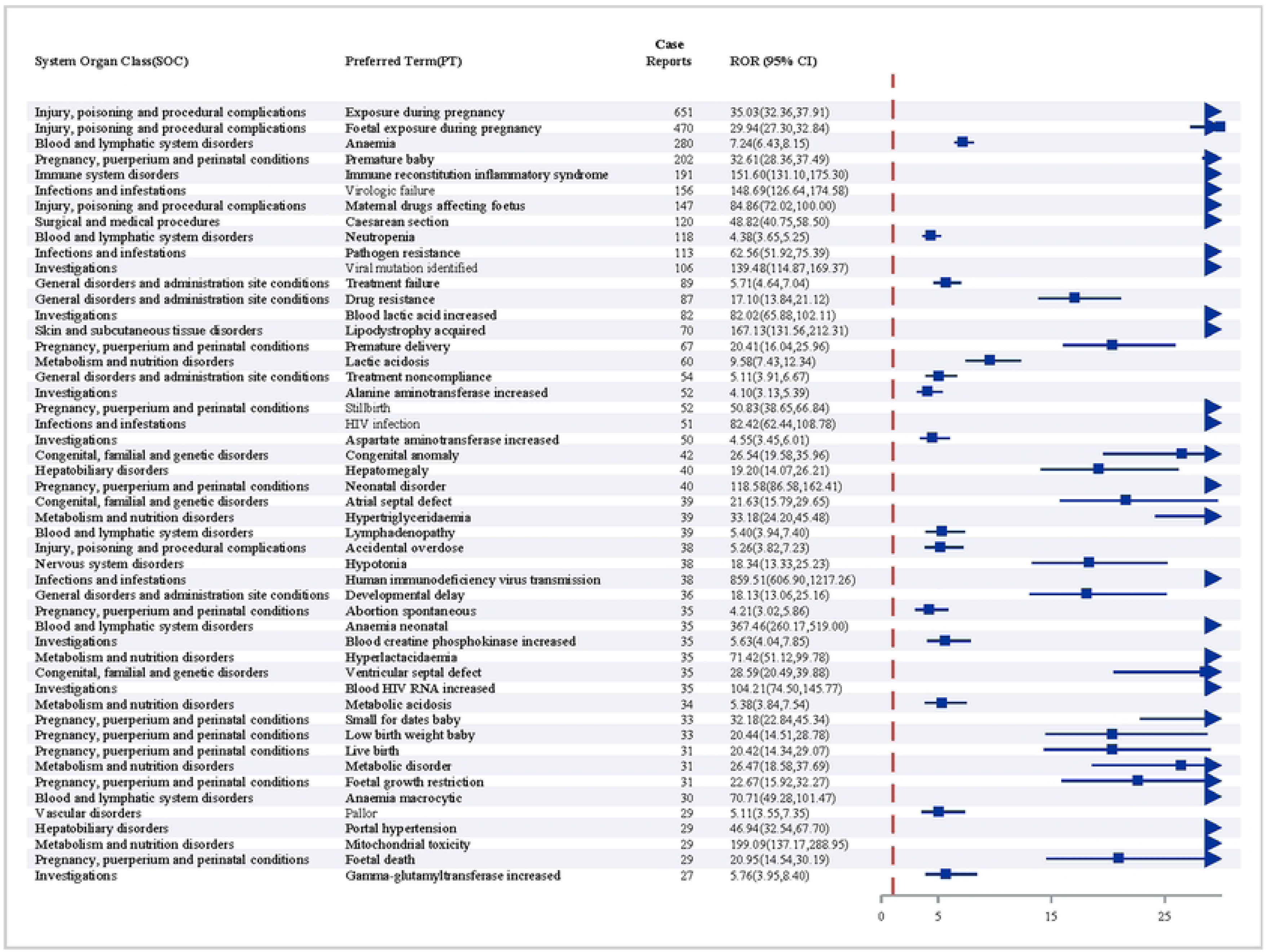
Signal strength of adverse events at the Preferred Term lev.

Within the pregnancy and congenital disorder subsets, key preferred terms with significant signals were identified, each corresponding to distinct clinical events without repetition: Premature baby [n = 202, ROR (95% CI) = 32.61 (28.36, 37.49)], Maternal drugs affecting foetus [n = 147, ROR (95% CI) = 84.86 (72.02, 100.00)], Caesarean section [n = 120, ROR (95% CI) = 48.82 (40.75, 58.50)], Premature delivery [n = 67, ROR (95% CI) = 20.41 (16.04, 25.96)], Stillbirth [n = 52, ROR (95% CI) = 50.83 (38.65, 66.84)], Congenital anomaly [n = 42, ROR (95% CI) = 26.54 (19.58, 35.96)], Neonatal disorder [n = 40, ROR (95% CI) = 118.58 (86.58, 162.41)], Spontaneous abortion [n = 35, ROR (95% CI) = 4.21 (3.02, 5.86)], Small for dates baby [n = 33, ROR (95% CI) = 32.18 (22.84, 45.34)], Low birth weight baby [n = 33, ROR (95% CI) = 20.44 (14.51, 28.78)], Foetal growth restriction [n = 31, ROR (95% CI) = 22.67 (15.92, 32.27)], and Foetal death [n = 29, ROR (95% CI) = 20.95 (14.54, 30.19)]. Each PT represents a unique adverse outcome with clear clinical distinctions. It is important to note that Caesarean section, while showing a statistical association, may be influenced by clinical decisions related to HIV management rather than directly attributable to zidovudine.

Additionally, we identified potential new AE signals not documented in the current product label, including acquired lipodystrophy, atrial septal defect, hypertriglyceridaemia, lymphadenopathy, hypotonia, ventricular septal defect, metabolic acidosis, portal hypertension, and mitochondrial toxicity. These signals, particularly those involving congenital structural anomalies (e.g., atrial septal defect, ventricular septal defect) and metabolic disorders, warrant further investigation in targeted studies involving pregnant populations.

### 3.3 Onset time of events

We extracted data on the onset time of zidovudine-related adverse events from the FAERS database, focusing on reports with documented timing of event occurrence relative to the initiation of zidovudine therapy. After rigorous screening to exclude records with incomplete or ambiguous timing information, a total of 509 AEs with valid onset time data were identified.

The median time to onset of zidovudine-related AEs was 23.00 days (interquartile range [IQR]: Q1 0.00, Q3 168.00), as depicted in Fig 4. This indicates that, on average, AEs associated with zidovudine tend to occur within the first month of treatment initiation, though there is considerable variability in the timing of onset.

**Fig 4.**
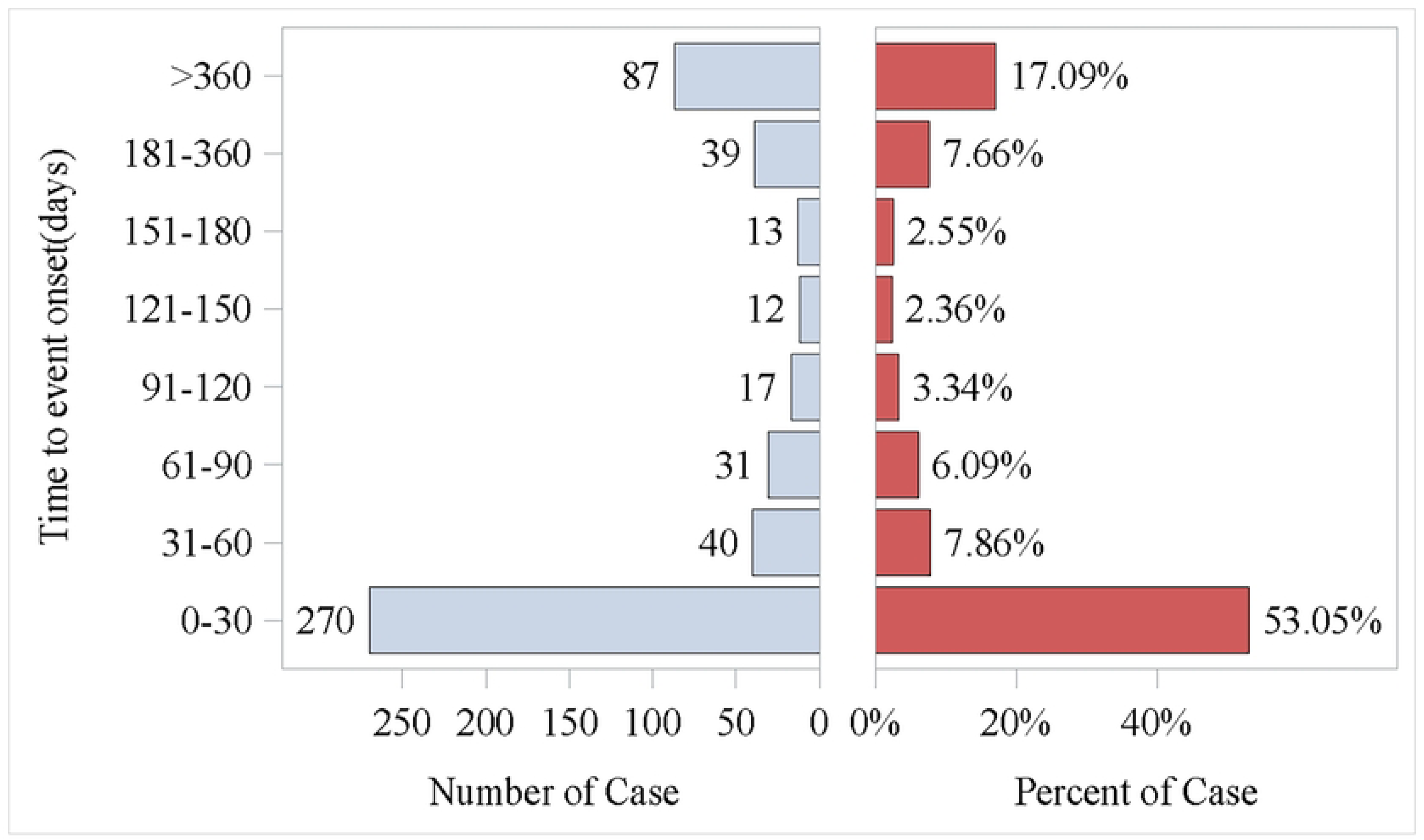
Time to event report distribution of AE reports.

A notable proportion of cases (n = 270, 53.05%) occurred within the first month following the start of zidovudine therapy. This early onset pattern suggests that close monitoring during the initial stages of treatment is crucial, regardless of the patient population. The proportion of cases reported in subsequent months showed a gradual decline: 40 cases (7.86%) in the second month, 31 cases (6.09%) in the third month, and 17 cases (3.34%) in the fourth month. This trend indicates that the risk of AEs decreases progressively over the first few months of treatment.

Additionally, the data revealed that 87 AEs (17.09% of the total) occurred after one year of zidovudine treatment. This long-term onset subgroup is important to consider, as it may reflect cumulative effects of prolonged zidovudine exposure or delayed reactions that manifest over extended periods of use. However, it is important to note that without stratification by patient population, we cannot determine the extent to which these long-term cases are associated with specific groups, such as non-pregnant individuals who may receive zidovudine for chronic therapy, or pregnant individuals where such long-term use is less typical.

In summary, the onset time of zidovudine-related AEs is characterized by a predominance of early occurrences within the first month, followed by a gradual decline in risk over the subsequent months, with a smaller but notable proportion of events occurring after one year of treatment. These findings emphasize the need for ongoing monitoring throughout the duration of zidovudine therapy, with particular attention to the initial months of treatment when the risk of AEs is highest.

## 4. Discussion

### 4.1 Principal Findings

In this study, we investigated the statistical associations between zidovudine and adverse pregnancy outcomes, as well as congenital disorders, at the SOC, SMQ, and PT levels. Leveraging a substantial dataset from the FAERS and employing multiple disproportionality analysis methods (ROR, PRR, BCPNN, and MGPS), our results demonstrated statistically significant associations between zidovudine and adverse events related to both adverse pregnancy outcomes and congenital disorders across all three levels of analysis.

The study included 2,931 patients receiving zidovudine, with 12,586 associated AE reports. Notably, the proportions of AE reports among patients under 18 years of age (17.13%) and those aged 18 to 44 years (17.33%) were nearly identical. According to Joint United Nations Programme on HIV/AIDS(UNAIDS), as of 2023, HIV-positive individuals aged 15 years and older account for over 90% of total infections, while less than 5% occur in those younger than 15 years old(19). This discrepancy suggests that despite the smaller proportion of younger patients utilizing zidovudine, the rate of reported AEs is disproportionately high in this age group. This finding is clinically relevant given that numerous prior studies have demonstrated that a considerable number of adverse drug reactions (ADRs) emerge during the first few years of life(20–23). For instance, to reduce perinatal HIV transmission, infants born to HIV-positive mothers are recommended to receive antiretroviral therapy (ART) as soon as possible after birth, ideally within 6 hours of delivery(24), and zidovudine is frequently administered to newborns for this purpose(25). Consequently, clinicians must remain vigilant regarding zidovudine-related AEs in younger patients, as early identification is critical to preventing life-threatening effects or disease progression.

Consistent with zidovudine ’ s established indications, the reported AEs were predominantly associated with prophylaxis and treatment of HIV. Geographically, the United States accounted for the highest percentage of reports (27.16%), followed by France (13.78%), with a peak in reporting observed in 2016. This surge may correlate with key studies published around that time: a 2016 U.S. study examining prenatal zidovudine exposure and its association with defect-free adverse birth outcomes(9), and a 2015 French study investigating associations between in utero zidovudine exposure and congenital heart defects (CHDs), as well as long-term myocardial remodeling in female infants postpartum(26). hese studies likely heightened clinical and public health concerns regarding zidovudine-related AEs, potentially contributing to increased reporting.

Disproportionality analyses identified significant signals for pregnancy and congenital disorders at the SOC level, including "Pregnancy, puerperium and perinatal conditions" and "Congenital, familial and genetic disorders"—both of which met the threshold criteria for all four analytical methods. At the SMQ level, relevant reports were categorized under "Complications and Risk Factors of Pregnancy, Labor, and Delivery (other than Miscarriage and Stillbirth)," "Congenital, Familial, and Hereditary Diseases," and "Termination of pregnancy and risk of abortion." These findings align with a systematic review and meta-analysis, which found that zidovudine was associated with an increased risk of perinatal complications compared to alternative nucleoside reverse transcriptase inhibitors among HIV-positive pregnant women(8), reinforcing the relevance of our observations to current clinical practice.

### 4.2 Results

The World Health Organization (WHO) currently recommends tenofovir disoproxil fumarate in combination with lamivudine or emtricitabine as first-line treatment for HIV-positive pregnant women(27). Zidovudine, abacavir, or tenofovir alafenamide, used in conjunction with lamivudine or emtricitabine, are listed as alternative options. Notably, zidovudine ’ s demotion from first-line status is attributed primarily to procedural considerations rather than safety concerns(28), though our findings—alongside prior research—add nuance to this distinction. Specifically, a systematic review and meta-analysis evaluating adverse perinatal outcomes in HIV-positive pregnant women receiving various NRTIs found that zidovudine was linked to a higher risk of adverse outcomes compared to other NRTIs(8), aligning with our pharmacovigilance (PV) signals.

Our identification of atrial septal defect as a PV signal suggests potential cardiotoxicity associated with zidovudine, which resonates with earlier studies documenting specific associations between in utero zidovudine exposure and congenital heart defects(29, 30). Mechanistic studies provide support for this link: cellular investigations indicate that zidovudine induces concentration-dependent cardiomyocyte death, characterized by activation of caspases-3 and -7, poly (ADP-ribose) polymerase, and increased mitochondrial reactive oxygen species (ROS) production in human cardiomyocytes(31, 32). Animal experiments further demonstrate that zidovudine-treated mice develop dose-dependent cardiac dilatation, impaired function, and ventricular inflammatory infiltrates, alongside elevated myocardial expression of Fas/Fas ligands, enhanced caspase-3 activation, calpain 1 translocation, and increased apoptosis(33–35). These preclinical findings reinforce the biological plausibility of our clinical observations.

Mitochondrial dysfunction emerges as a recurring theme in zidovudine ’ s toxicity profile. Clinical studies of placentas from HIV-positive pregnant women exposed to combination antiretroviral therapy (cART) containing zidovudine show mitochondrial DNA (mtDNA) depletion, heightened oxidative stress, and increased apoptosis—indicators of secondary mitochondrial impairment that may contribute to adverse perinatal outcomes(36, 37). However, whether the mitochondrial toxicity represents a zidovudine-specific effect or is confounded by HIV infection and pregnancy physiology remains unresolved. One study indicates that elevated oxidative stress in HIV-positive pregnant women is linked to both the virus itself and gestational physiology(38),underscoring the need for further research to delineate zidovudine’ s independent contribution to this pathway. Notably, the pharmacovigilance signal we identified—zidovudine inhibition of mtDNA replication, leading to decreased synthesis of striated muscle mtDNA, mtRNA, and mitochondrial peptides (39–41)—aligns closely with prior mechanistic findings, collectively implicating mitochondrial disruption as a central mechanistic pathway.

Beyond cardiac and mitochondrial effects, our analysis identified PT signals linking zidovudine to metabolic disorders, including acquired lipodystrophy, hypertriglyceridaemia, and metabolic disorder.These findings are consistent with studies showing that zidovudine exposure in HIV-exposed but uninfected infants and children is associated with metabolic abnormalities(42), and in vitro research demonstrating that zidovudine inhibits cell proliferation while increasing lactate and lipid production(43, 44). Changes in adipocytokines have also been observed in human adipocyte cell lines and in vivo studies of NRTI-treated HIV patients, with zidovudine prominently implicated(45, 46). Additionally, our PT signals for blood lactic acid increased, lactic acidosis, and metabolic acidosis align with zidovudine ’ s labeled adverse effects(47, 48), while signals for anaemia, neonatal anaemia, and macrocytic anaemia—also documented in the product label—further validate the robustness of our PV approach.

It is important to contextualize these findings within broader ART strategies: several studies note that prenatal combination ART reduces early HIV transmission rates compared to zidovudine monotherapy but may carry a higher risk of adverse maternal and neonatal outcomes(49, 50). These findings collectively highlight the need for a balanced approach when selecting perinatal ART regimens, weighing the substantial benefits of transmission reduction against the potential risks of adverse outcomes. Our study, which focuses exclusively on zidovudine, does not address these comparative risks, underscoring the need for future research evaluating safety profiles across ART regimens in pregnancy.

### 4.3 Clinical Implications

Our findings inform clinical practice in optimizing zidovudine use among HIV-positive pregnant women. Targeted monitoring—including fetal growth assessments and cardiac imaging— should be prioritized, especially during the first month of treatment when AE risk is highest, aligning with WHO guidance on balancing efficacy and safety(28). Detection of new signals (e.g., acquired lipodystrophy, mitochondrial toxicity) highlights the need for comprehensive post-marketing surveillance, with metabolic monitoring integrated into follow-up care for mothers and exposed infants. Our data support the shift to tenofovir-based regimens(28), but in settings where zidovudine is necessary, clinicians should discuss potential risks (e.g., congenital anomalies) while emphasizing its role in preventing mother-to-child transmission. Proactive monitoring of hematological and metabolic parameters can facilitate timely intervention, mitigating severe outcomes in vulnerable populations.

In summary, our findings refine clinical decision-making, enhance surveillance, and improve maternal and fetal outcomes among HIV-positive pregnant women treated with zidovudine.

### 4.4 Research Implications

This study contributes to the field of pharmacovigilance by providing robust evidence of zidovudine-associated safety signals in pregnancy and congenital disorders using real-world data from FAERS. The identification of both known (e.g., anaemia, lactic acidosis) and novel (e.g., atrial septal defect, acquired lipodystrophy) signals underscores the value of post-marketing surveillance in complementing pre-approval clinical trials, which often underrepresent pregnant populations. These findings can inform the design of targeted studies—such as prospective cohort investigations of mitochondrial function in zidovudine-exposed fetuses or comparative effectiveness research on ART regimens in resource-limited settings—to further clarify causal relationships and mechanisms. Additionally, the methodological framework employed here (integrating multiple disproportionality metrics) offers a replicable model for evaluating drug safety in vulnerable populations, supporting evidence-based updates to global HIV treatment guidelines.

### 4.5 Strengths and Limitations

A key strength of this study is the large, multi-decade dataset from FAERS, which captures real-world AE reports across diverse geographic regions and clinical contexts, enhancing the generalizability of our findings. The use of four complementary disproportionality analysis methods (ROR, PRR, BCPNN, MGPS) also strengthens the robustness of identified safety signals, particularly for understudied outcomes like congenital heart defects in zidovudine-exposed infants.

Notably, this study has several limitations. First, as a spontaneous reporting system, FAERS is prone to underreporting and reporting bias, with variability in how AEs are interpreted and documented. For example, 55.34% (n=1,622) of cases lacked age information due to data recording errors, complicating the stratification of congenital disorder reports by age group—though some neonatal cases could be inferred from AE terms. Second, critical clinical details such as gestational age, stillbirth autopsy results, and concurrent ART medications are unavailable in FAERS, limiting our ability to isolate zidovudine-specific effects from confounding factors (e.g., HIV-related pathophysiology or co-administered drugs). Third, the cross-sectional nature of FAERS data prevents definitive causal inference, as temporal relationships between zidovudine exposure and AEs (e.g., long-term metabolic disorders) cannot be fully validated.

These limitations highlight the need for cautious interpretation of our findings, particularly when extrapolating to clinical decision-making. Integrating these results with prospective clinical data and individual patient factors (e.g., gestational age, pre-pregnancy ART use) remains essential for balanced risk-benefit assessments.

## 5. Conclusion

This pharmacovigilance study analyzed FAERS data (Q1 2004–Q4 2024) to assess zidovudine’s associations with adverse pregnancy outcomes and congenital disorders using four disproportionality methods. Significant signals were confirmed at SOC, SMQ, and PT levels, including "Pregnancy, puerperium and perinatal conditions," "Congenital, familial and genetic disorders," and specific outcomes like preterm birth and congenital anomalies. Novel unlabeled signals (e.g., acquired lipodystrophy, atrial septal defect) were also identified.

Findings highlight the value of post-marketing surveillance for zidovudine, particularly in pregnant populations where it remains critical for preventing mother-to-child HIV transmission. Early onset of most AEs (within 1 month) necessitates targeted monitoring, while long-term events require sustained vigilance in prolonged therapy.

Limitations include FAERS underreporting bias, missing clinical details, and inability to confirm causality. Future research should validate novel signals, clarify mechanisms (e.g., mitochondrial dysfunction), and compare ART regimens in pregnancy.

Overall, this study enhances understanding of zidovudine’s safety profile, aiding clinicians and regulators in optimizing use to improve maternal and fetal outcomes in HIV-positive populations.

## Data Availability

All relevant data are within the manuscript and its Supporting Information files.

## Abbreviations

ADRs: Adverse drug reactions
AEs: Adverse Events
APR: Antiretroviral Pregnancy Registry
ART: Antiretroviral therapy
ARV: Antiretroviral
AZT: Zidovudine
BCPNN: Bayesian Confidence Propagation Neural Network
cART: Combined antiretroviral therapy
EBGM: Empirical Bayesian Geometric Mean
FAERS: FDA Adverse Event Reporting System
FDA: US Food and Drug Administration
HEU: HIV-exposed uninfected
HIV: Human immunodeficiency virus
LBW: low birth weight
MedDRA: Medical Dictionary for Regulatory Activities
MGPS: Multi-item Gamma Poisson Shrinker
mtDNA: Mitochondrial DNA
NRTI: Nucleoside reverse transcriptase inhibitor
NTS: Nucleus of the solitary tract
PRR: Proportional reporting ratio
PS: Primary suspect
PT: Preferred term
PV: Pharmacovigilance
ROR: Reporting odds ratio
ROS: Reactive oxygen species
SMQs: Standardised MedDRA queries
SOC: System organ class
UNAIDS: Joint United Nations Programme on HIV/AIDS
WHO: World Health Organization
WLHIV: Women living with HIV

## Author contributions

Zhongxiang Zhang: Data curation, Methodology, Resources, Software, Visualization, Writing– original draft, Writing–review and editing. Xinyun Du: Data curation, Methodology, Writing– original draft, Writing–review and editing. Liuyi Ren: Conceptualization, Methodology, Supervision, Visualization, Writing–original draft. Long He: Data curation, Methodology, Supervision, Visualization. Xuping Yang: Conceptualization, Methodology, Software, Supervision. Qiaoying Li: Software,Visualization, Writing–original draft. Kun Tu: Software,Visualization. Shurong Wang: Data curation, Methodology, Resources, Visualization, Writing–review and editing. Jie Zhou: Data curation, Methodology, Resources, Software, Visualization, Writing–original draft, Writing–review and editing. Yilan Huang: Data curation, Methodology, Resources, Software, Visualization, Writing– original draft, Writing–review and editing.

## Acknowledgements

Thanks to the US FDA for providing the data source of this study.

## Data availability statement

The original contributions presented in the study are included in the article/Supplementary Material, further inquiries can be directed to the corresponding authors.

## Supporting Information

**S1 table.**
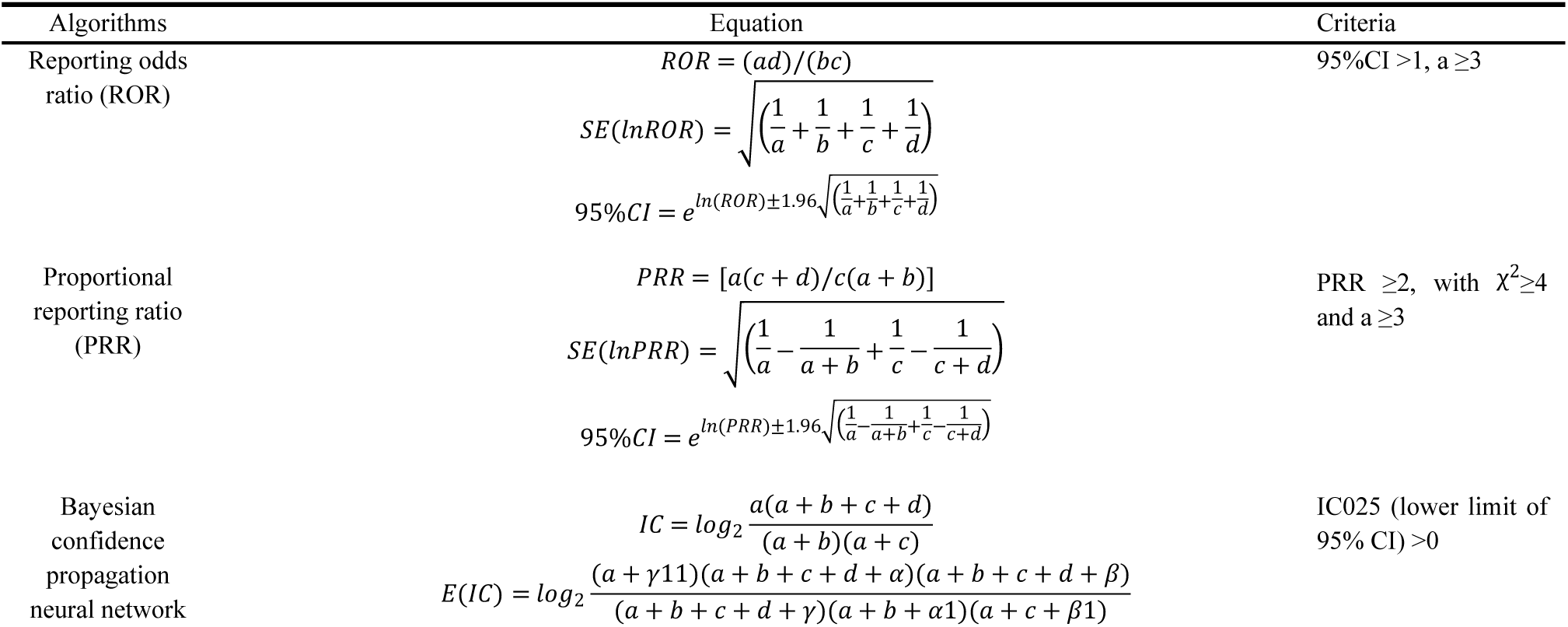
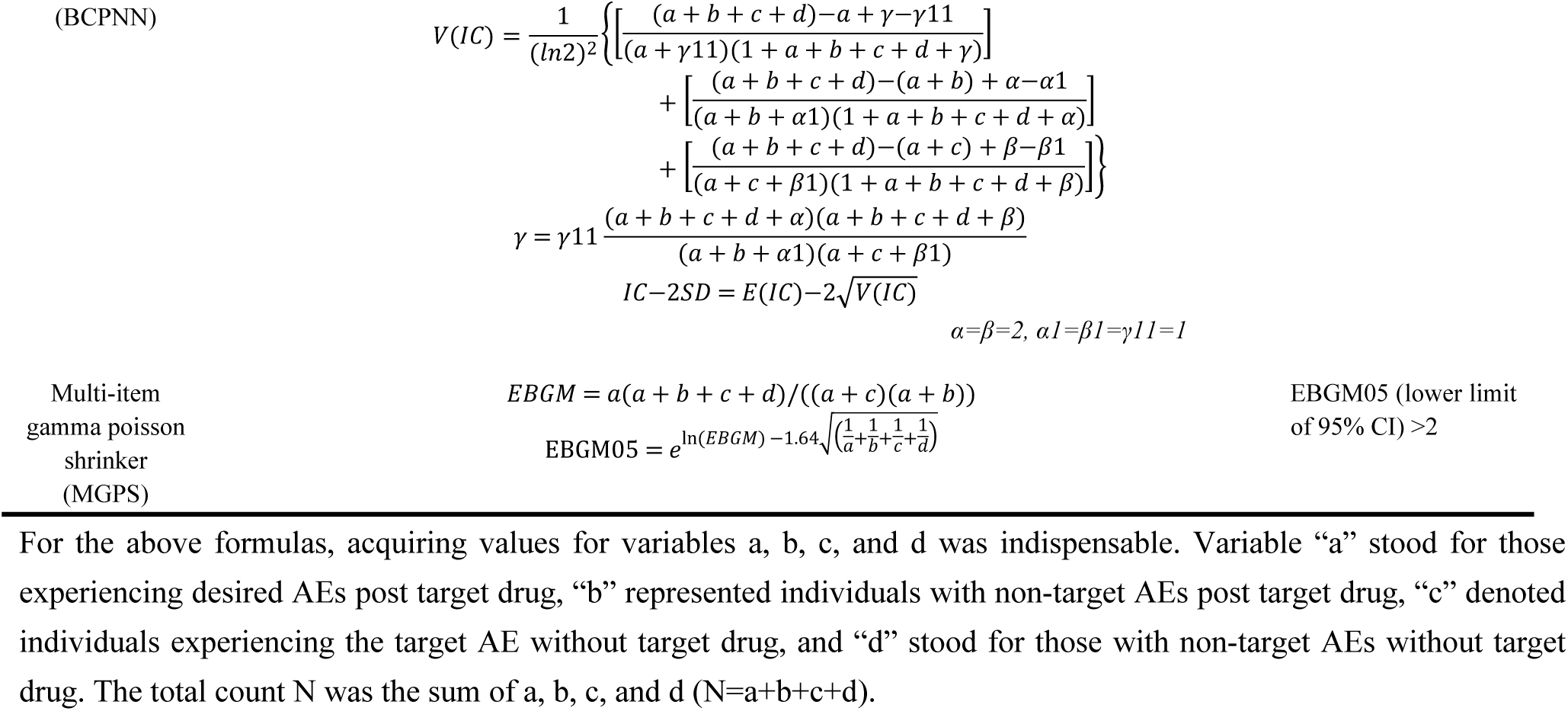
Four methods of disproportionality analysis.

**S2 table.**
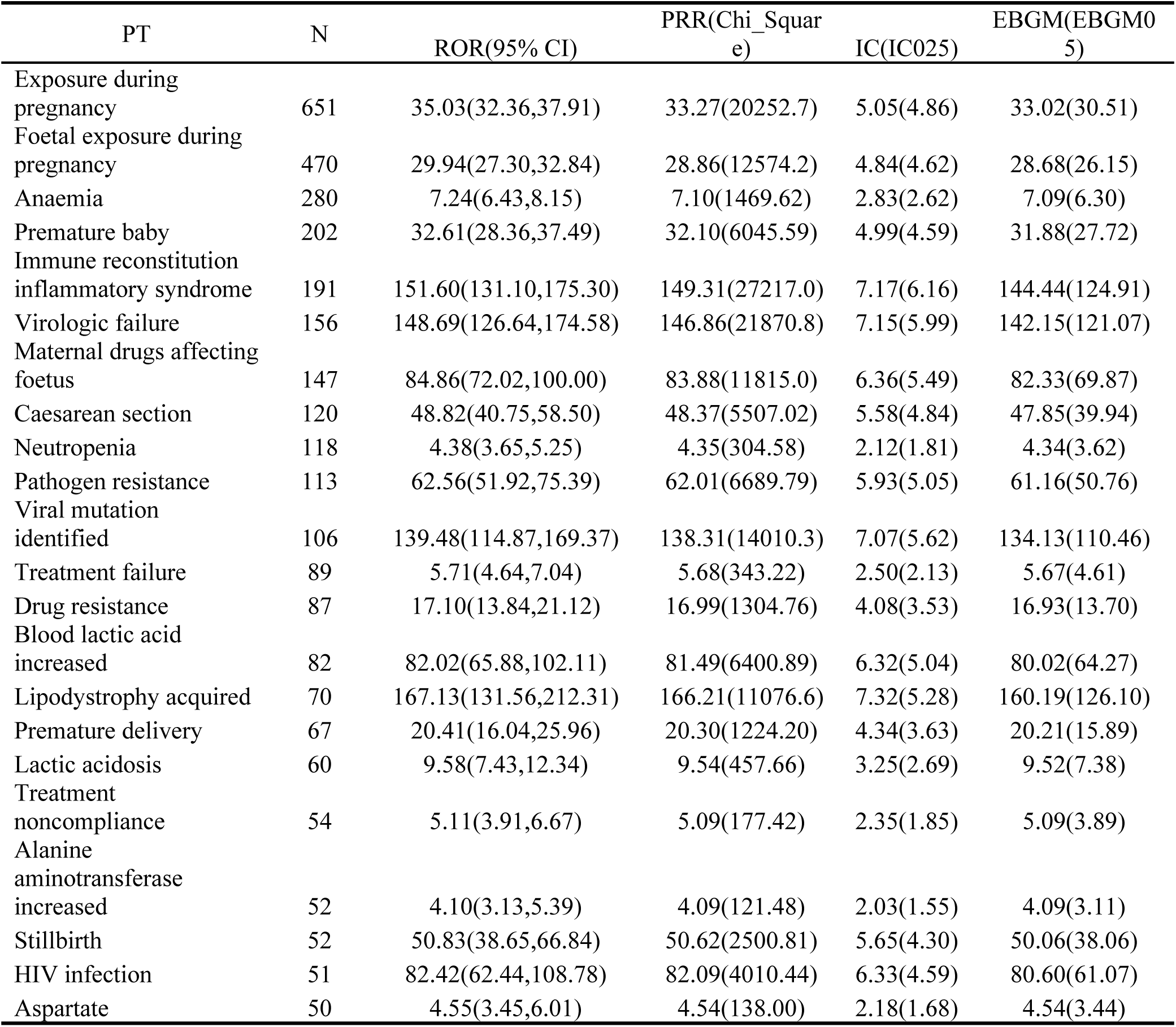
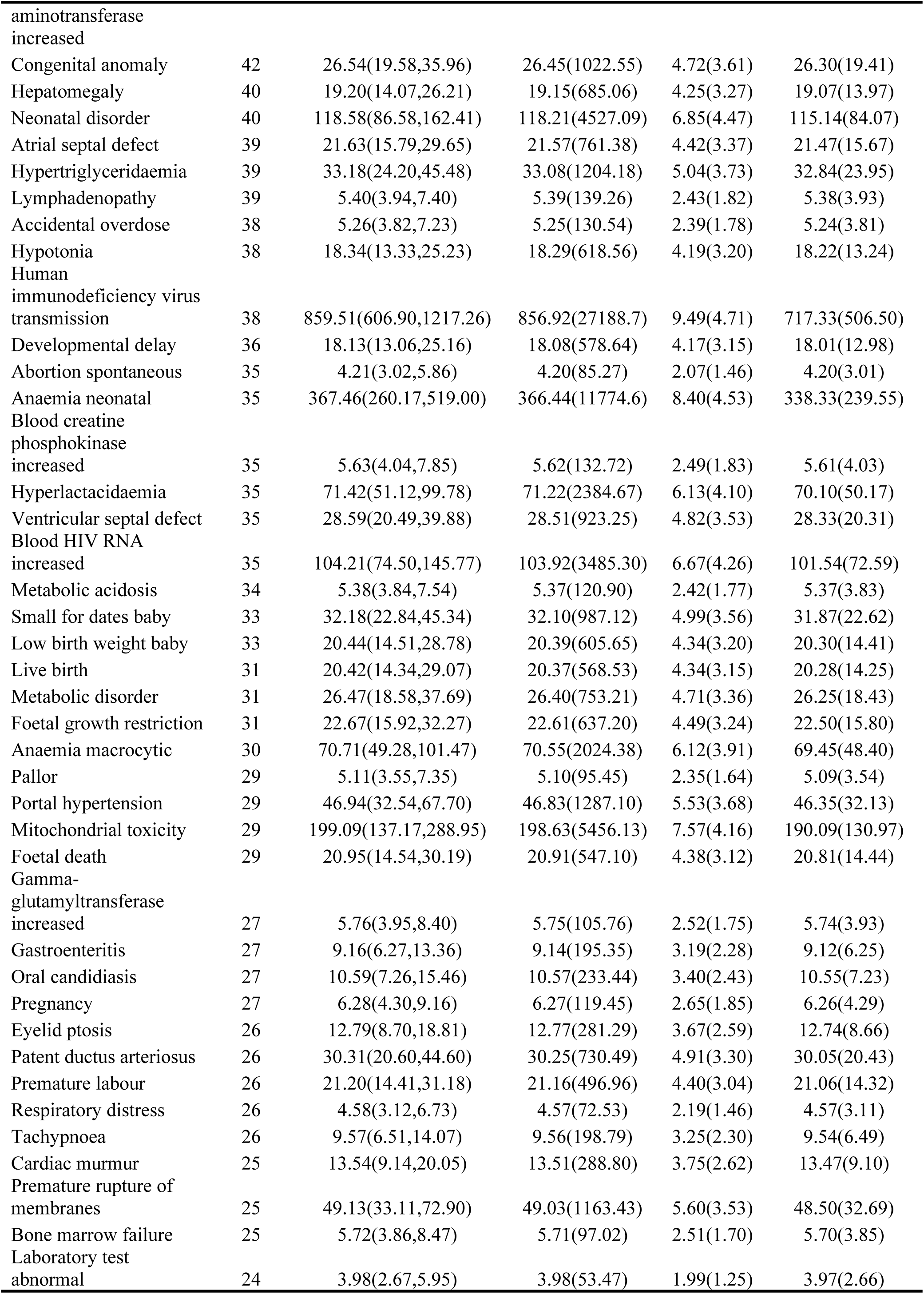
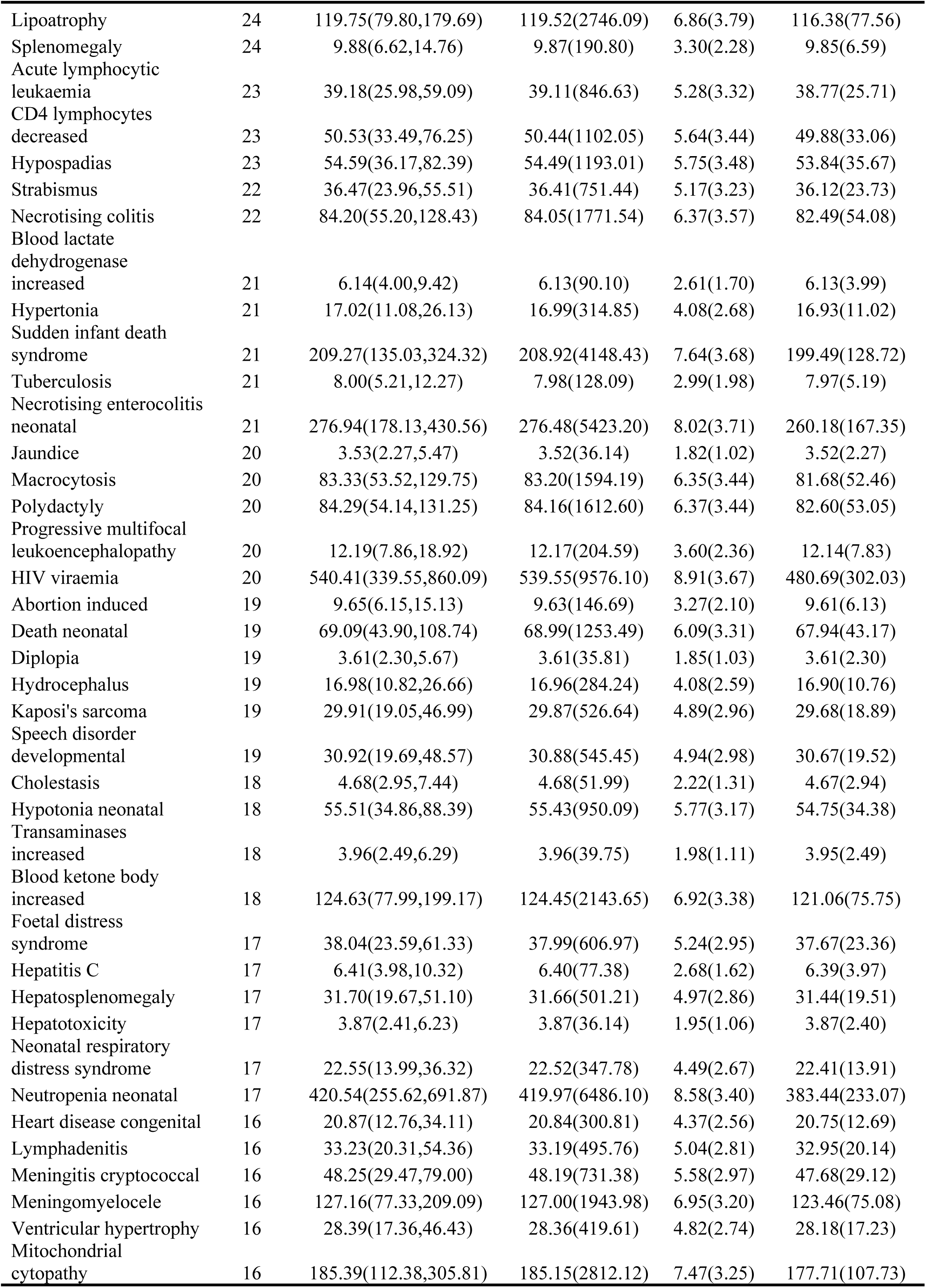
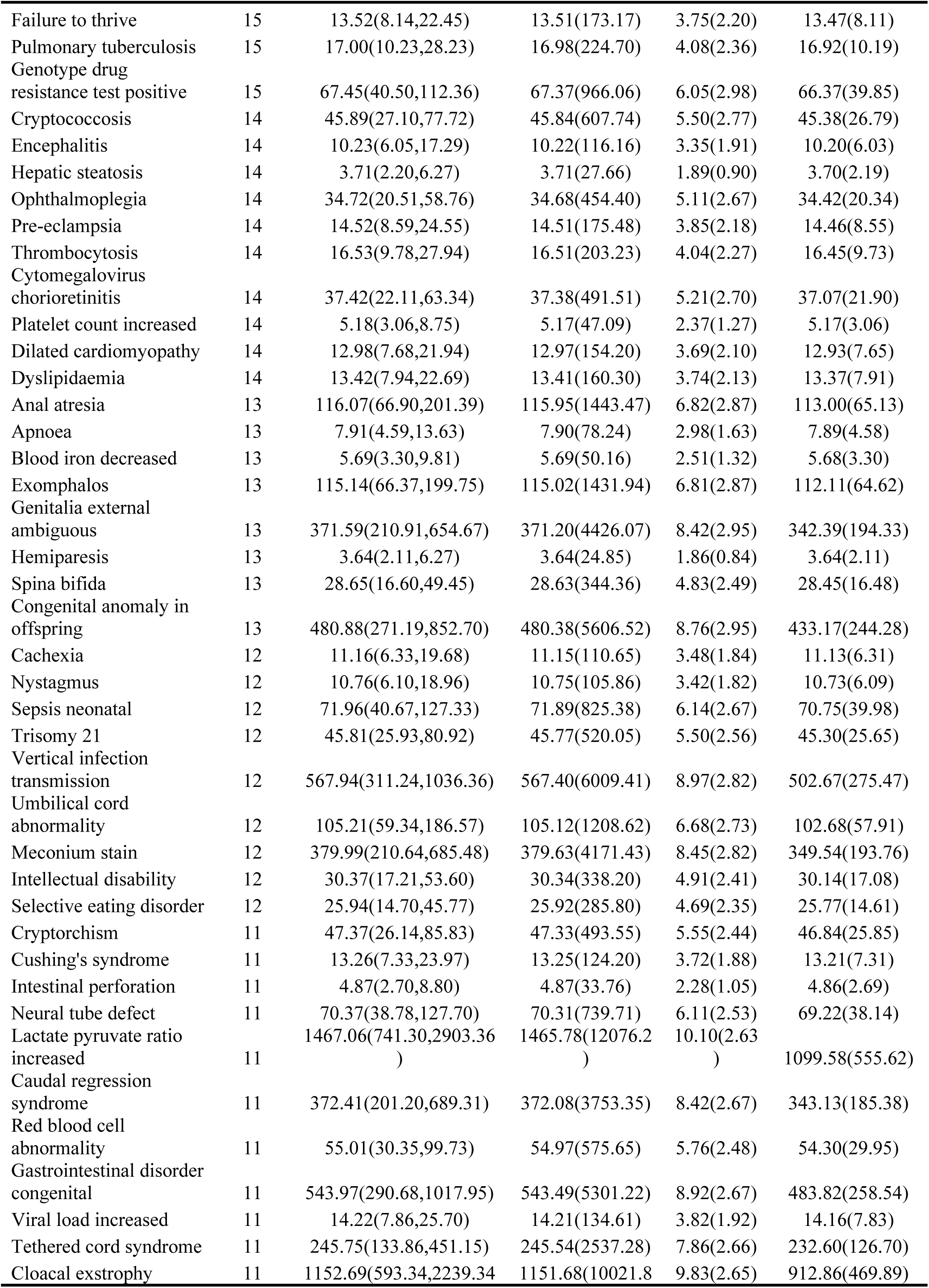
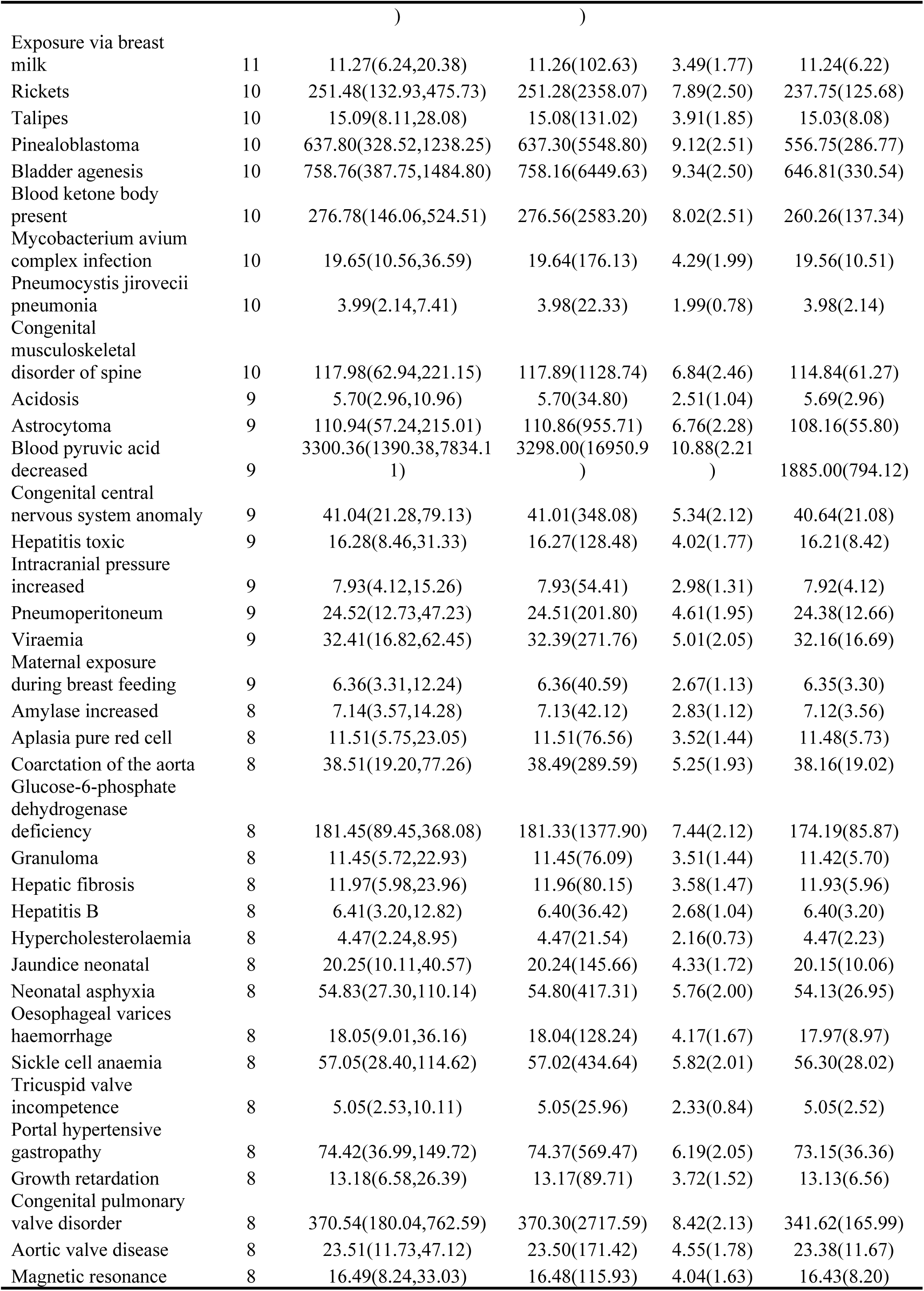
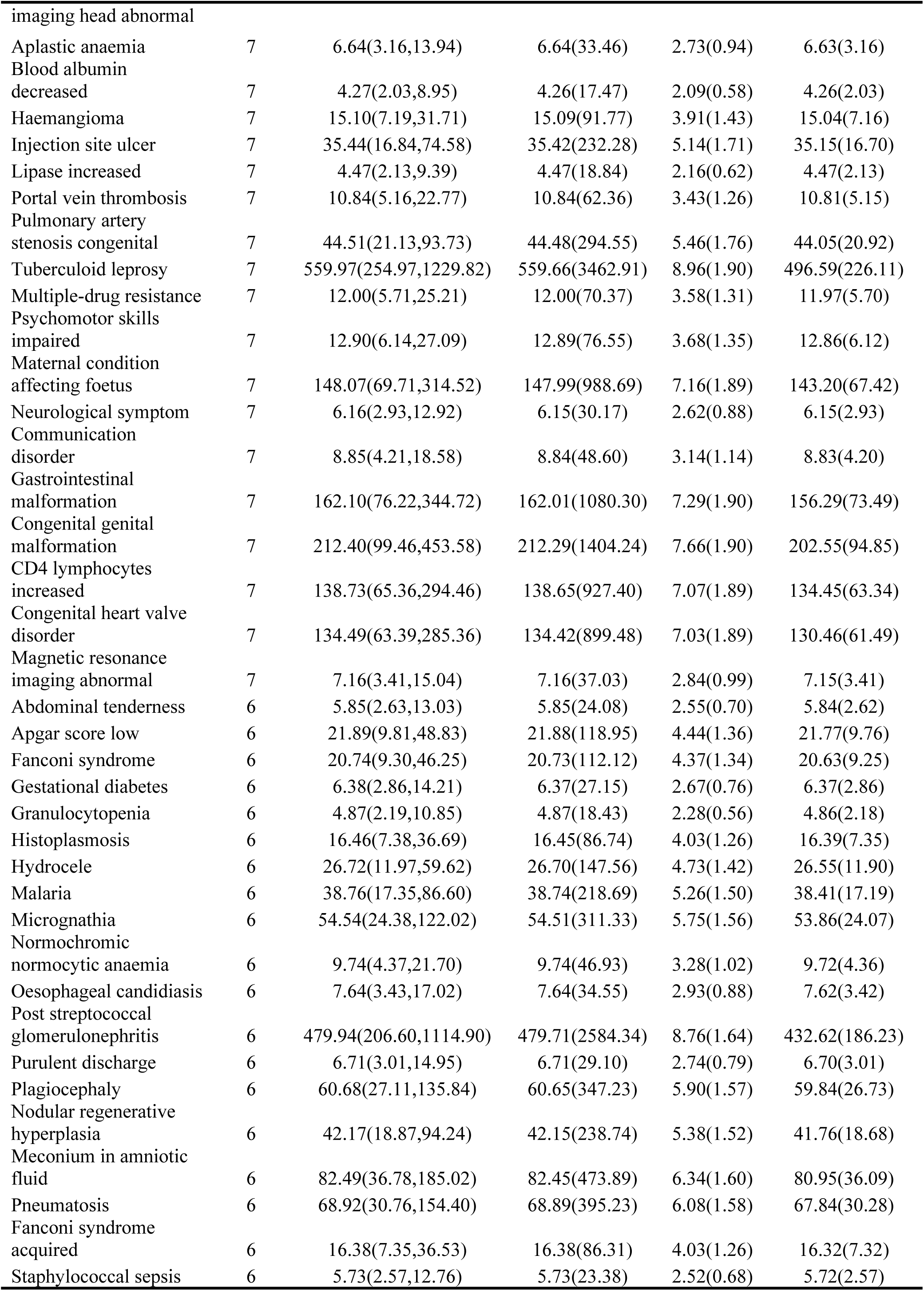
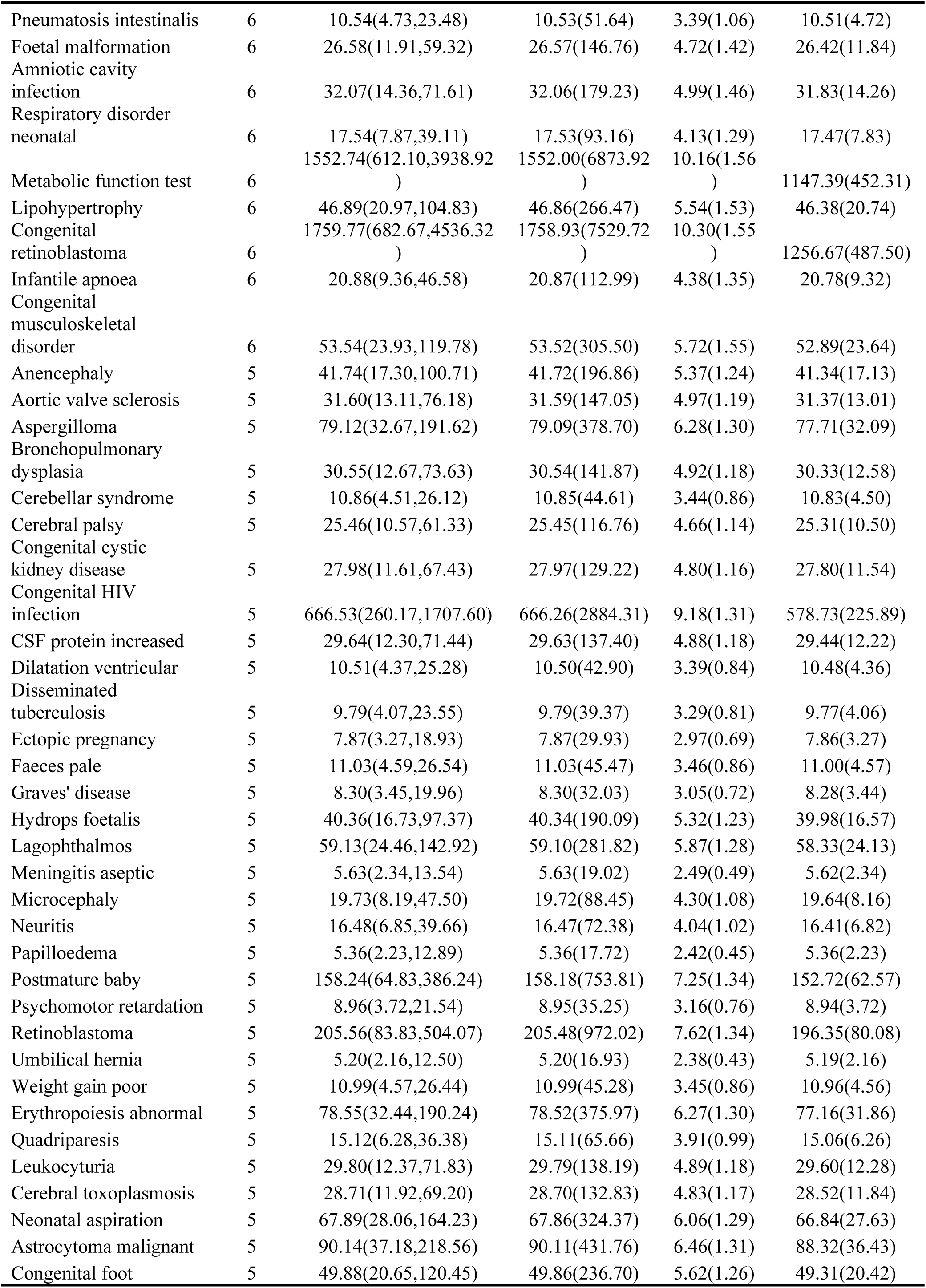
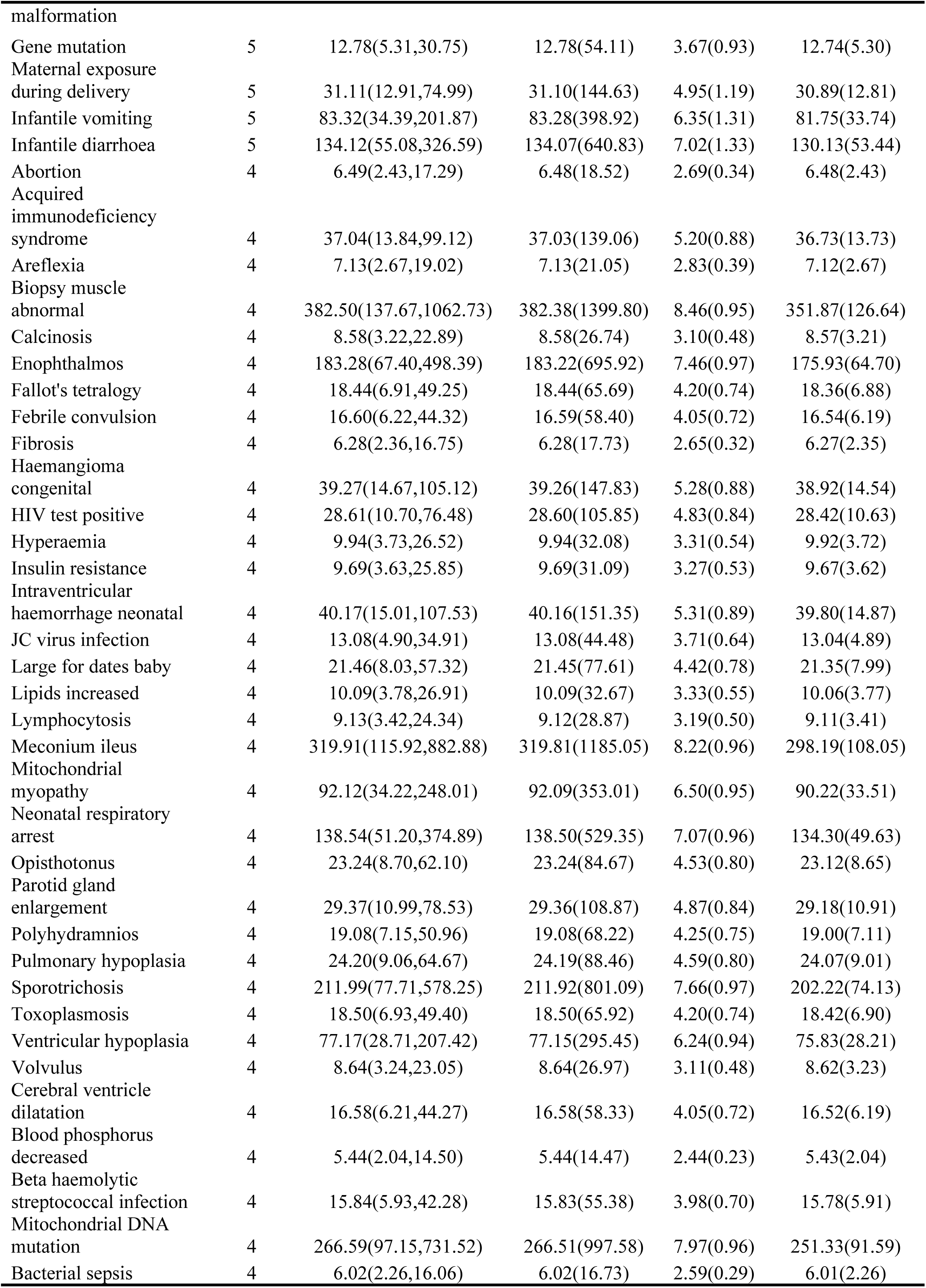
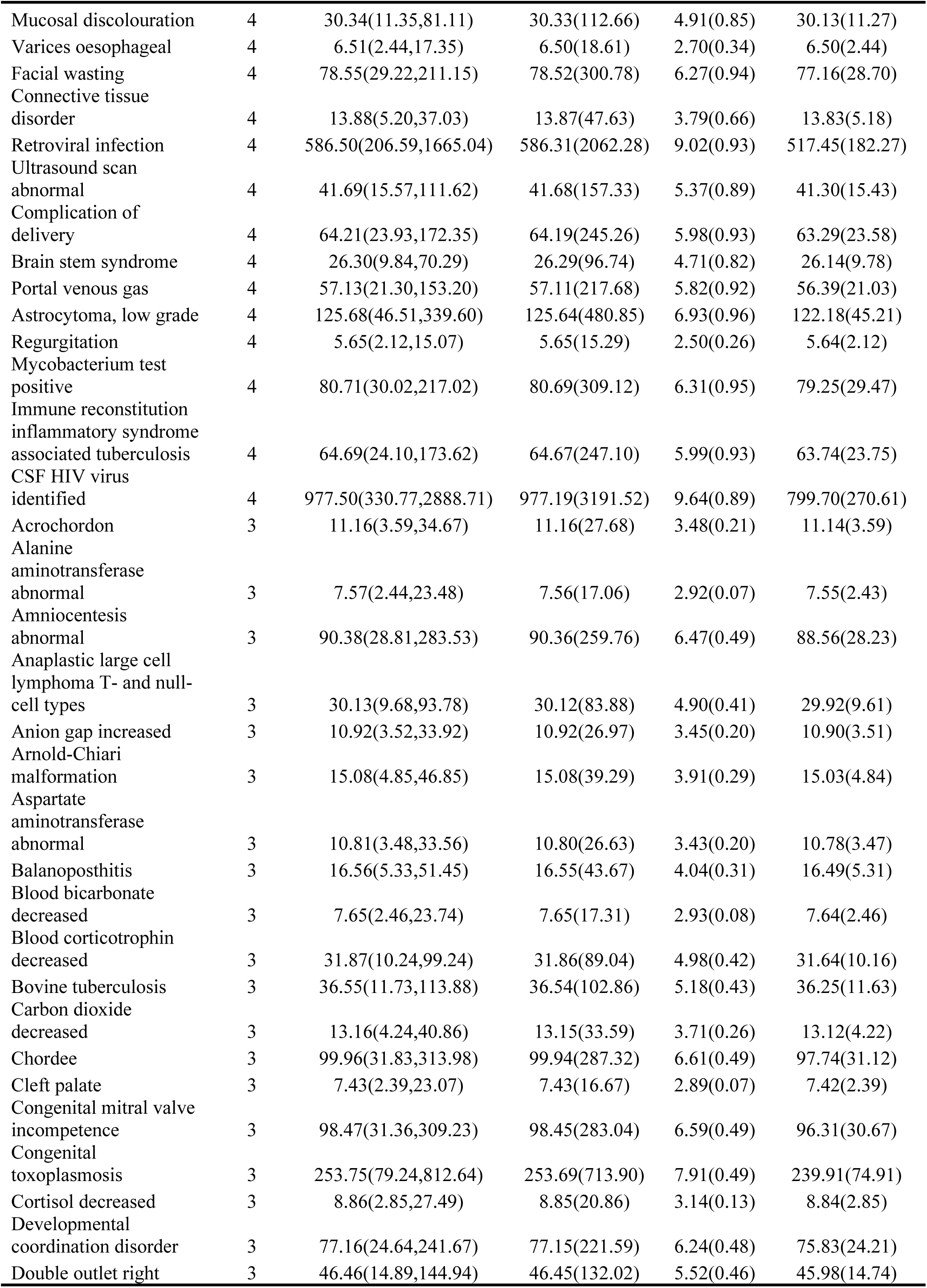
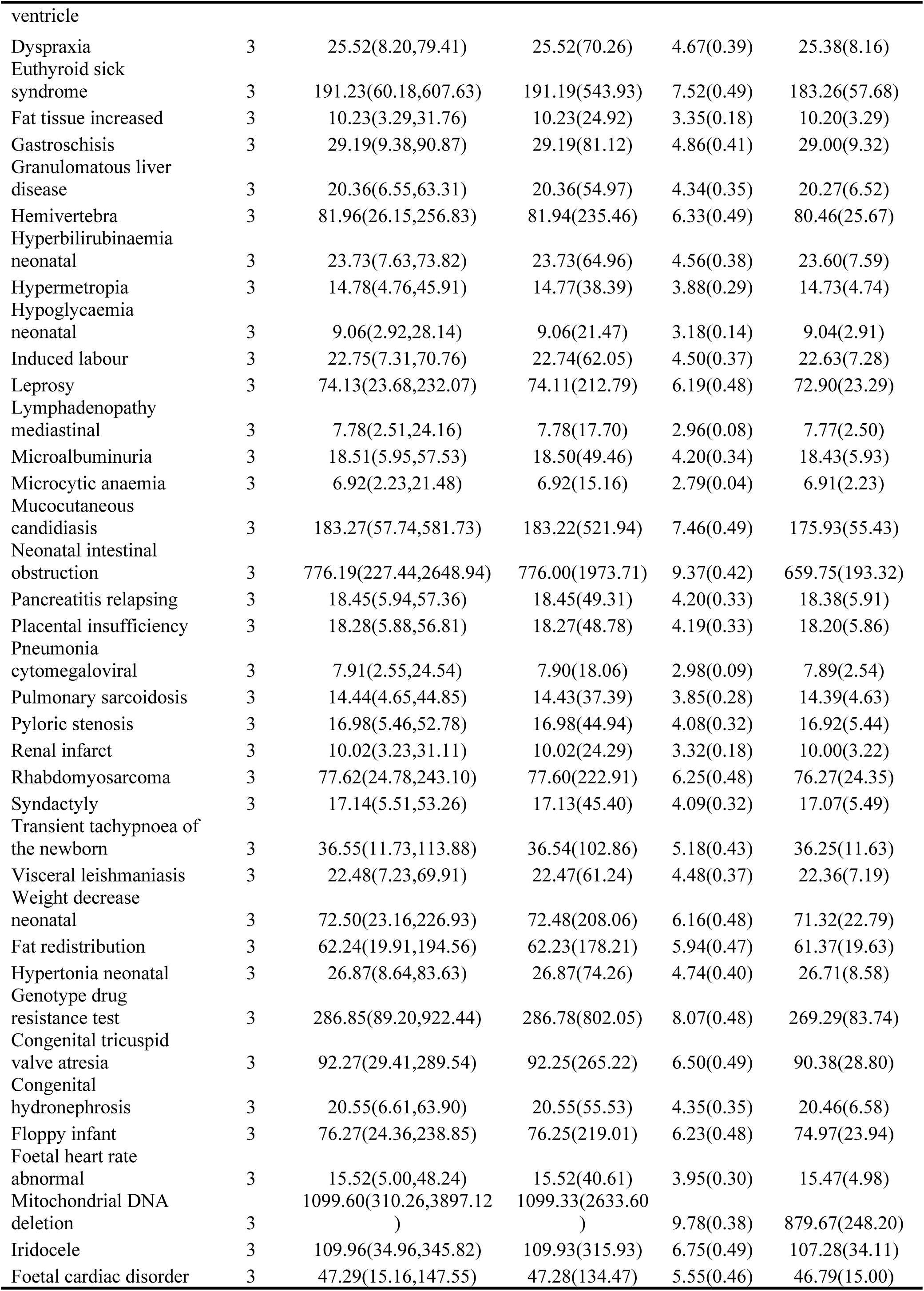
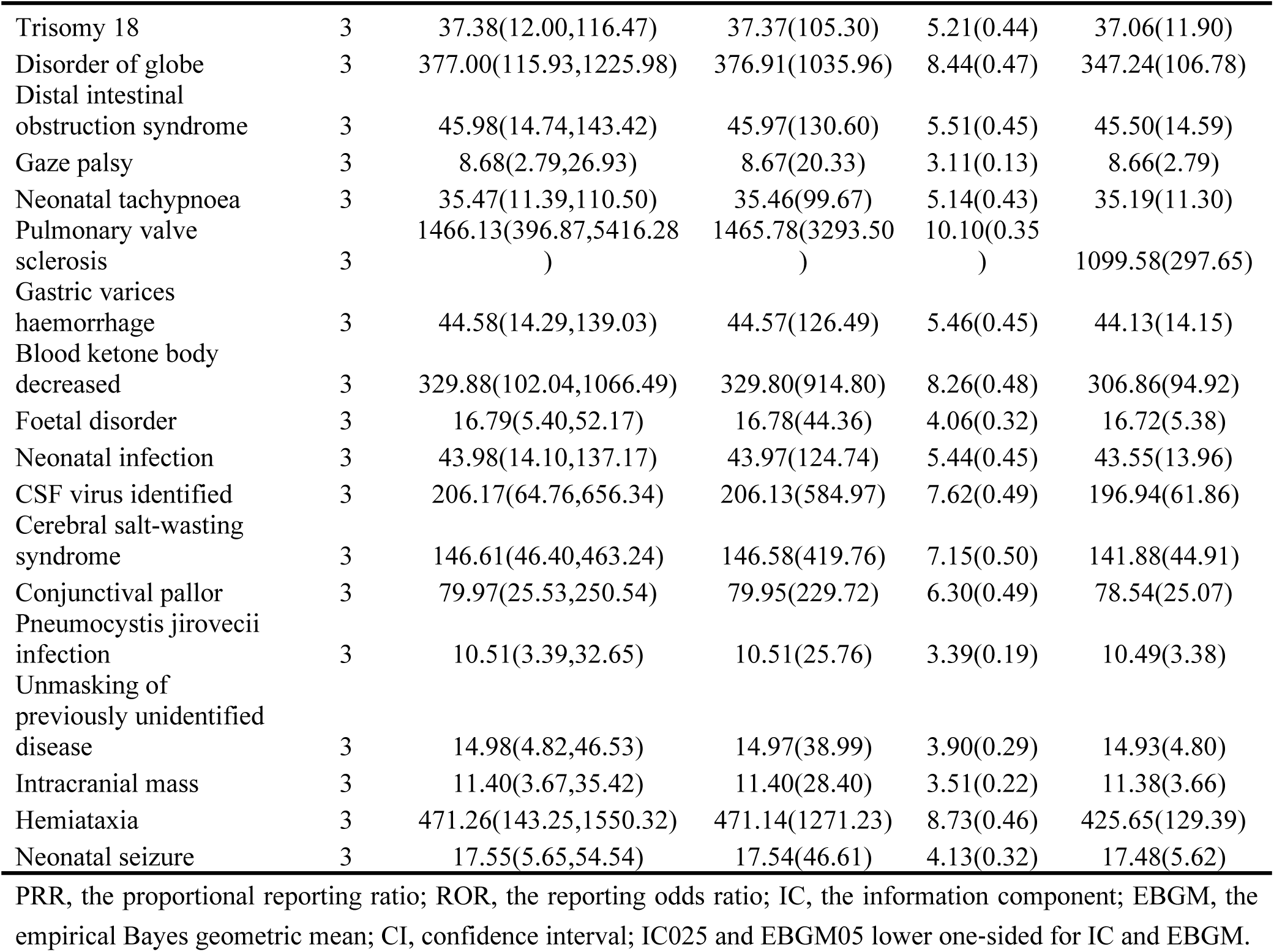
Disproportionality analysis of signals at the PT level.

